# Equilibrium properties of a coupled contagion model of mosquito-borne disease and mosquito preventive behaviors

**DOI:** 10.1101/2025.02.01.25321519

**Authors:** Marya L. Poterek, Mauricio Santos-Vega, T. Alex Perkins

## Abstract

Although different strategies for mosquito-borne disease prevention can vary significantly in their efficacy and scale of implementation, they all require that individuals comply with their use. Despite this, human behavior is rarely considered in mathematical models of mosquito-borne diseases. Here, we sought to address that gap by establishing general expectations for how different behavioral stimuli and forms of mosquito prevention shape the equilibrium prevalence of disease. To accomplish this, we developed a coupled contagion model tailored to the epidemiology of dengue and preventive behaviors relevant to it. Under our model’s parameterization, we found that mosquito biting was the most important driver of behavior uptake. In contrast, encounters with individuals experiencing disease or engaging in preventive behaviors themselves had a smaller influence on behavior uptake. The relative influence of these three stimuli reflected the relative frequency with which individuals encountered them. We also found that two distinct forms of mosquito prevention—namely, personal protection and mosquito density reduction—mediated different influences of behavior on equilibrium disease prevalence. Our results highlight that unique features of coupled contagion models can arise in disease systems with distinct biological features.

## 1 Introduction

Dengue virus is a mosquito-borne pathogen transmitted by *Aedes aegypti* and *Ae. albopictus* mosquitoes that poses a risk to approximately half of the world’s population [1]. Currently, there is no treatment for dengue and only one moderately effective vaccine available, so interventions that target mosquitoes are the primary means of dengue prevention [2]. These interventions span a variety of specific techniques, but they often boil down to either large-scale insecticide spraying, typically performed by government agencies, or in-home water container management, typically performed by residents and reinforced with educational campaigns [3, 4]. Empirical studies suggest that “intersectoral” approaches combining spraying and community-driven control are more successful than spraying or community control strategies alone, although it is difficult to generalize across settings [5].

An important factor in the success of either of these strategies is human behavior, since interventions can only be effective if they are adopted in the first place. Behavioral choices can influence compliance with spraying campaigns and participation in mosquito larval habitat reduction, sometimes producing unexpected outcomes. For example, outdoor spatial spraying alone has been associated with a lower adoption of in-home water container management and higher entomological risk, as observation of outdoor spatial spraying can give a false sense of security [6, 7, 8]. Inclusion of educational programming in a campaign can counteract this effect, however [9]. Most importantly, though, clinical trials of community interventions have reported up to a 25% reduction in Aedes-borne diseases [10, 11], demonstrating that individual behavior can have a measurable impact on transmission.

Mathematical modeling has been used to elucidate the interplay between disease and behavioral dynamics for a variety of directly transmitted pathogens, particularly those impacted by vaccine hesitancy [12, 13] and changing contact patterns [14, 15, 16]. Forecasting models that account for individual behavioral change can produce significantly different forecasts than those that disregard adaptive behaviors [16], which could compromise forecast accuracy [17, 18]. There are comparatively few published models of mosquito-borne disease dynamics that include behavior, and much remains unknown about the relationships among mosquito density, disease prevalence, and preventive behaviors. Previous studies have explored relatively narrow questions around this topic, such as how changing mosquito preventive behaviors in the presence of a dengue vaccine and varying intervention effectiveness, can impact transmission [19, 20]. Those studies have also provided insight into the impacts of information sharing across spatiotemporal scales and targeted public health messaging on mosquito-borne disease incidence [21, 22].

When disease-related behavior itself is conceptualized as an infectious entity, a “coupled contagion” model presents a useful framework for understanding the feedback between such behaviors and disease. This approach to modeling cocirculating contagious processes together, rather than independently, specifically considers how transmission is impacted as the contagions evolve and interact. For directly transmitted pathogens, reducing contact via social distancing or isolation are the primary forms of preventive behavior. Several studies coupling adaptive behaviors and directly transmitted pathogens found that even limited amounts of fear-driven self-isolation can drive multiple waves of infection in an epidemic scenario [23, 24, 25, 26, 27], while targeted public health messaging strategies could minimize outbreak size across a communication network [28]. Mosquito-borne diseases, in contrast, can be influenced by a wider range of preventive behaviors. In addition to reducing the mosquito biting rate through the use of a repellent, other preventive behaviors for mosquito-borne diseases include those with an indirect effect on transmission, such as actions taken to reduce mosquito density. Likewise, mosquito-borne diseases are unique in that exposures to mosquito biting could prompt individuals to engage in preventive behaviors, independent of disease status.

In this study, we sought to establish fundamental principles for the coupled-contagion dynamics of a mosquito-borne disease and mosquito prevention behaviors. Our model allowed for two distinct mosquito prevention behaviors: use of personal protection (which only confers direct protection) and reduction of mosquito larval habitat (which confers indirect protection by reducing mosquito density). We considered three distinct influences that would prompt individuals to engage in these behaviors: encountering people experiencing disease, encountering people engaging in preventive behaviors, and encountering mosquitoes. Because our model with fully coupled dynamics of disease and behavior was not analytically tractable, we first performed separate analyses of the equilibria of each component model. We then explored the equilibria of the coupled model using numerical simulations, with a focus on understanding how the assumptions and parameters underpinning the uptake and impact of preventive behaviors affect the equilibrium prevalence of behavior and, ultimately, disease.

## 2 Methods

### 2.1 Single contagion models

To explore the interface between mosquito-borne disease and mosquito preventive behaviors, we developed two deterministic compartmental models for the standalone dynamics of disease and behavior. We established a constant human population size *N*,

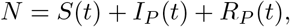

such that *N* ≡1, and leveraged this to characterize both single-contagion systems using analytical methods. We identified local equilibria using Mathematica 14.0 [29], and performed local stability analyses of those equilibria using the caracas R package (*version 2*.*0*.*1*) [30].

#### 2.1.1 Disease model

We developed a susceptible-infected-recovered-susceptible (SIRS) model to describe disease dynamics, modeled after dengue. In reality, dengue viruses comprise four distinct serotypes that confer lifelong homologous immunity and temporary heterologous immunity [31]. Although the SIRS model we used does not capture the full complexity of these dynamics, it does capture the fact that waning heterologous immunity (as individuals transition from *R* to *S*) allows persistence of the four serotypes in aggregate. In this model, susceptible individuals *S* are subject to the force of infection for the pathogen, *βI*, such that

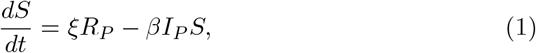

where the *P* subscript denotes that this model refers to infection with the pathogen. Infected individuals *I*_*P*_ recover from dengue at rate *γ* and lose immunity to dengue at rate *ξ*, or

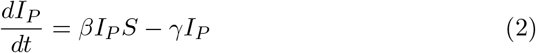

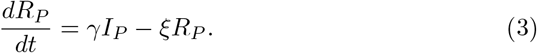

Given that 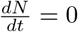 and *N ≡*1, we reduced this system to

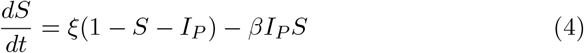

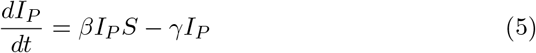

and performed our analyses thereon.

#### 2.1.2 Behavior model

We used a susceptible-infected-susceptible (SIS) model to describe the transmission of mosquito preventive behavior. In the absence of disease, susceptible individuals *S* become infected with the preventive behavior at a rate λ_*B*_ = ϕ _*M*_ *M* + ϕ _*S*_*I*_*B*_, so that

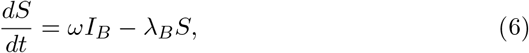

where the *B* subscript denotes that this model refers to infection with the behavior. Adoption of behavior occurs at a rate equal to the sum of the current ratio of mosquitoes to humans *M* and the proportion of individuals already performing the behavior *I*_*B*_, weighted by two parameters *ϕ* _*M*_ and *ϕ* _*S*_, respectively. The ratio of mosquitoes to humans varies over time as the proportion of the population engaged in preventive behaviors does, such that *M* (*t*) = *m*(1 − (1 − *α*_*M*_) *I*_*B*_), where 1 − *α*_*M*_ represents the efficacy of the preventive behavior in reducing mosquito density. Infected individuals then “recover” from this behavior at rate *ω*,

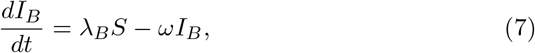

at which time they revert back to the state of no longer engaging in the behavior. We conducted the same analysis of this two-dimensional system as was done for our disease model.

### 2.2 Coupled contagion model

Using our understanding of single contagion model equilibria and parameter relationships, we integrated the two models into a single ODE system capturing the joint transmission of mosquito-borne disease and mosquito preventive behavior (Figure 1).

**Figure 1:**
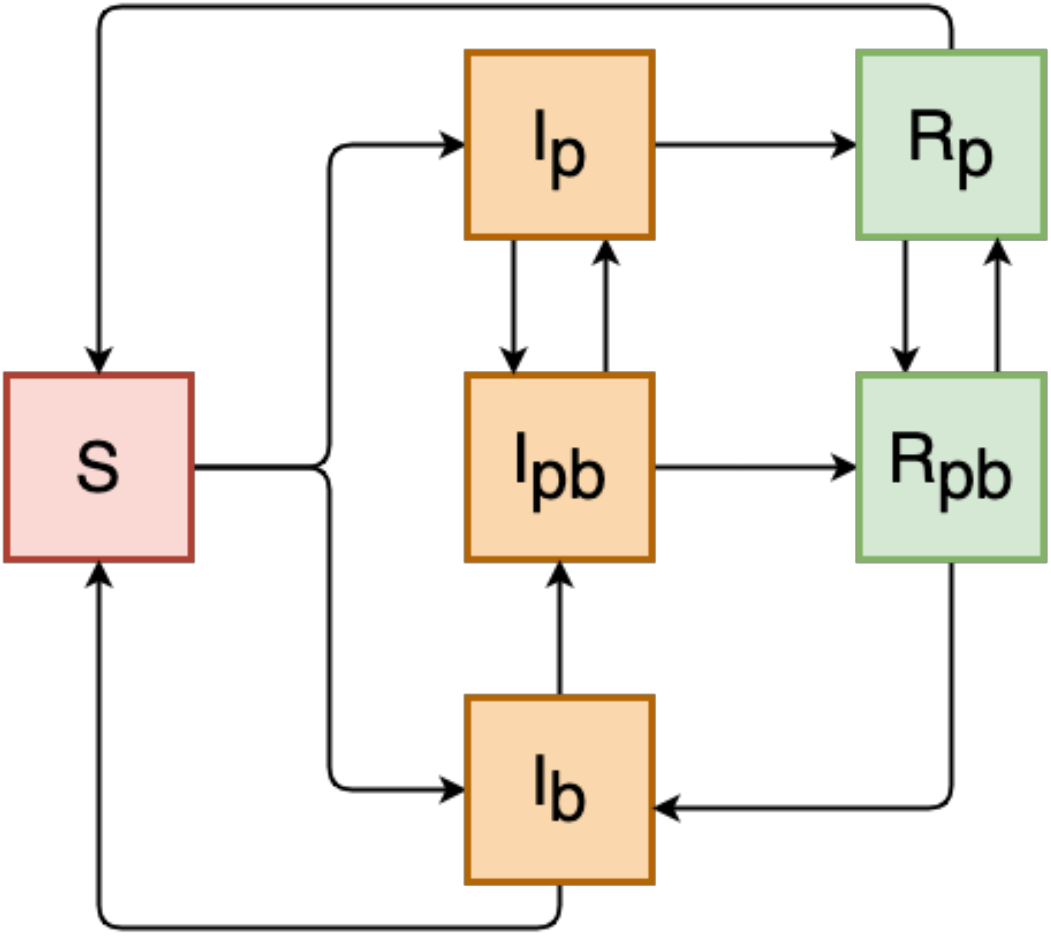
Diagram of coupled contagion model for dengue and behavior transmission. Compartment labels refer to susceptible *S*, infected *I*, and recovered *R* classes, while subscripts indicate the contagion associated with each state: *P* for the pathogen alone, *B* for behavior alone, and *PB* for both pathogen and behavior.

Here, the coupled dynamics of disease and behavior were modeled as an SIRS-SIS system in which humans can be infected with behavior alone (denoted by subscript *B*), pathogen alone (denoted by subscript *P*), or co-infected with both pathogen and behavior (subscript *PB*). Simultaneous co-transmission of pathogen and behavior is not allowed under our model, given that it would further complicate the model and is likely to be an exceedingly rare event. The coupled nature of the model allows for dynamic feedback between disease and behavior, which can interact in ways that drive long-term disease dynamics [25]. As before, we assumed a constant human population without demographic change, disease-induced mortality, or serotype dynamics. We summarize human population attributes as follows, where *N* represents the entire population, *N*_*B*_ the proportion of the population engaged in preventive behaviors, and *N*_*P*_ the proportion of the population infected with the pathogen, implying

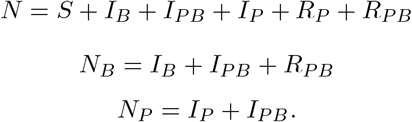

Susceptible individuals (*S*) in the model are naive to both pathogen and behavior, and become infected with the pathogen at rate λ_*P*_ and behavior at rate λ_*B*_. Those infected with the pathogen (*I*_*P*_) or behavior (*I*_*B*_) alone can then become co-infected (*I*_*PB*_) at rate λ_*B*_ or λ_*PB*_, respectively. Once infected, immunity to the pathogen and the behavior each wane at independent rates. System dynamics follow

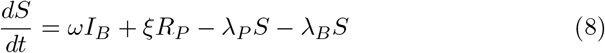

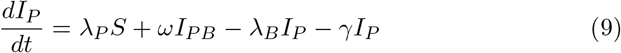

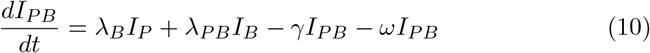

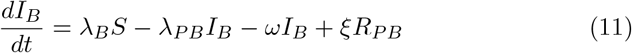

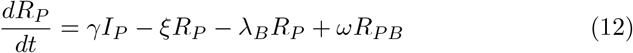

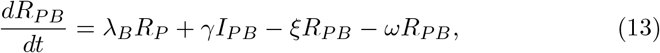

which is structurally consistent with our simpler, single contagion models.

We modified elements of both pathogen and behavior transmission to explicitly incorporate a mechanism for feedback between them. To do so, we first defined the force of infection for behavior, λ_*B*_, to include the number of individuals infected with the pathogen *N*_*P*_ (i.e., *I*_*P*_ + *I*_*PB*_), so that transmission of behavior is driven by a weighted sum of the proportion of the population infected with the pathogen, the adult mosquito prevalence, and the proportion of the population already engaged in the behavior, equal to

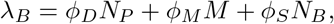

where *ϕ* _*D*_, *ϕ* _*M*_, and *ϕ* _*S*_ are the weights for these three respective behavioral stimuli. We chose these three stimuli because they span the range of possible influences on mosquito preventive behavior uptake [32, 33, 34].

The coupled model also features an expanded definition of the pathogen transmission rate *β* to include key components of mosquito biology and pathogen transmission [35]. By substituting the Ross-Macdonald model’s basic reproduction number expression for mosquito-borne pathogen transmission into our disease-only model’s *R*_0_ term, we can express the transmission rate as

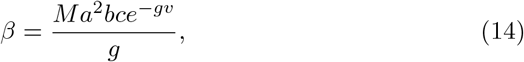

where *M* is the ratio of mosquitoes to humans, *a* is the blood-feeding rate, *b* is the probability that an infectious mosquito infects a susceptible human during blood-feeding, *c* is the probability that an infectious human infects a susceptible mosquito during blood-feeding, *g* is the mosquito mortality rate, and *v* is the extrinsic incubation period (Table 1). As in the behavior-only model, *M* is proportional to the population engaging in preventive behaviors, meaning that

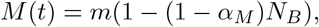

where 1 − *α*_*M*_ represents the reduction in mosquito density associated with preventive behaviors that result in mosquito larval habitat reduction [10].

**Table 1:**
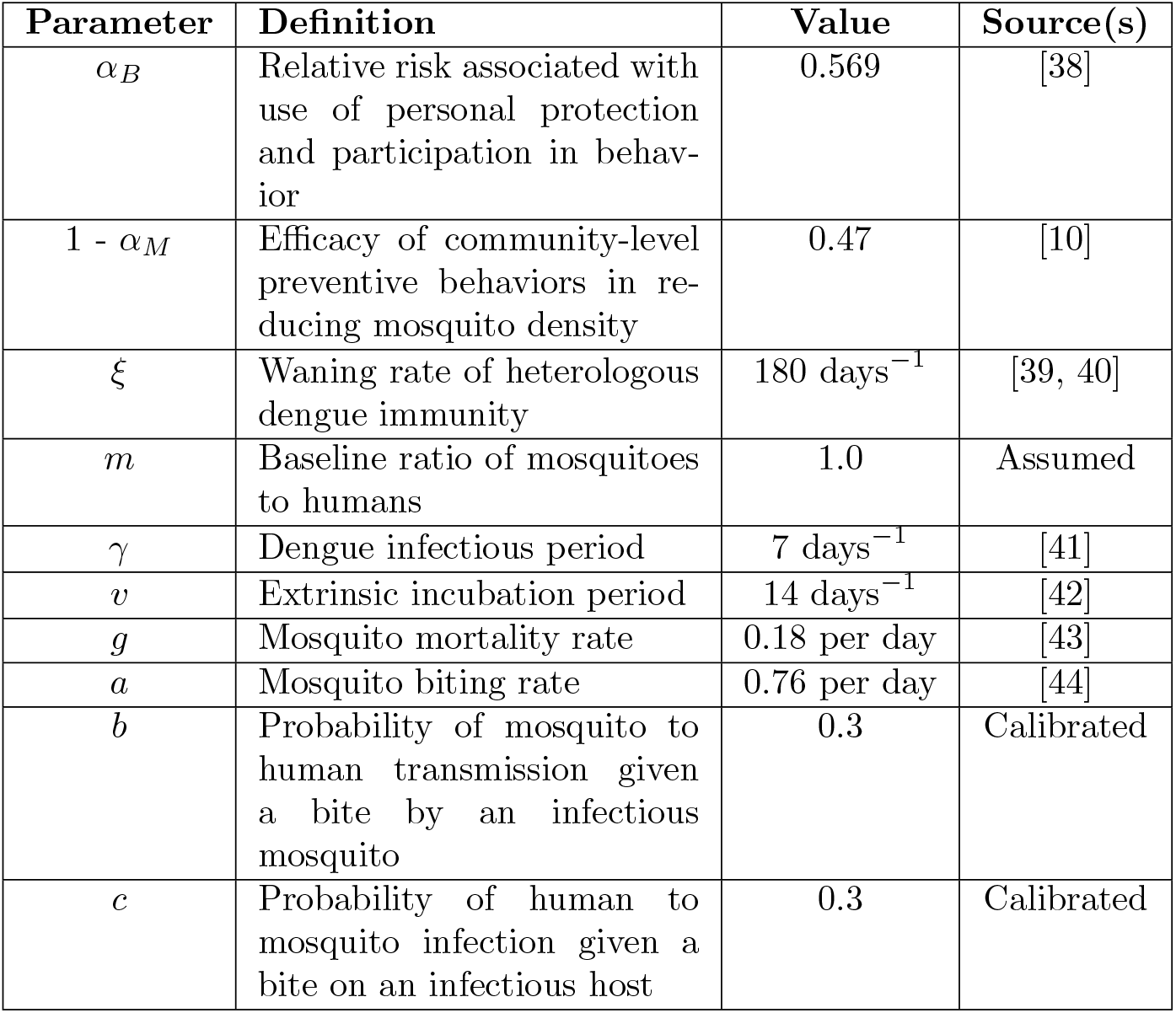
Disease parameters, definitions, and values. Parameters described as calibrated were set to match published estimates of *R*_0_ and force of infection for dengue virus [36, 37].

| Parameter | Definition | Value | Source(s) |
| --- | --- | --- | --- |
| $\alpha_B$ | Relative risk associated with use of personal protection and participation in behavior | 0.569 | [38] |
| $1 - \alpha_M$ | Efficacy of community-level preventive behaviors in reducing mosquito density | 0.47 | [10] |
| $\xi$ | Waning rate of heterologous dengue immunity | 180 days<sup>-1</sup> | [39, 40] |
| $m$ | Baseline ratio of mosquitoes to humans | 1.0 | Assumed |
| $\gamma$ | Dengue infectious period | 7 days<sup>-1</sup> | [41] |
| $v$ | Extrinsic incubation period | 14 days<sup>-1</sup> | [42] |
| $g$ | Mosquito mortality rate | 0.18 per day | [43] |
| $a$ | Mosquito biting rate | 0.76 per day | [44] |
| $b$ | Probability of mosquito to human transmission given a bite by an infectious mosquito | 0.3 | Calibrated |
| $c$ | Probability of human to mosquito infection given a bite on an infectious host | 0.3 | Calibrated |

In addition to these community-level effects on mosquito density, we assumed that individuals engaged in preventive behaviors also experienced some amount of personal protection through actions such as the use of mosquito repellent. The parameter *α*_*B*_ encapsulates this as a relative risk of infection. This shows up in our model as a squared effect given that it modifies the blood-feeding rate, *a*, and mosquitoes blood-feed twice in a transmission cycle (Equation 14). Thus, the corresponding forces of infection for those not performing and performing the behavior, respectively, are λ_*P*_ = *βN*_*P*_ and 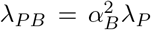.All behavioral parameters, definitions, and values are defined in Table 2.

**Table 2:**
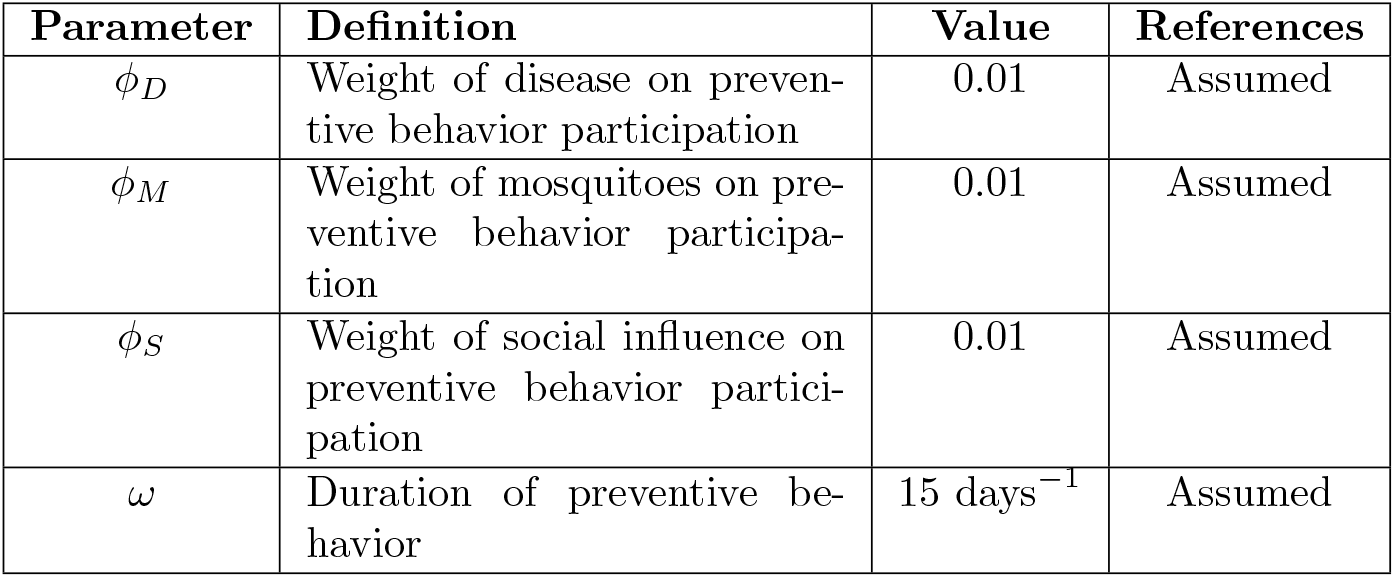
Behavioral parameters, definitions, and values.

We used numerical analyses to approximate model equilibria based on the prevalence attained after five years of model simulation, under a range of behavioral conditions. Simulations were performed using R Statistical Software (*version 4*.*3*.*2, 2023-10-31*) and the deSolve R package (*version 1*.*38*) [45, 46].

## 3 Results

### 3.1 Single contagion equilibria

#### 3.1.2 Disease model

This model possesses two equilibrium solutions: a disease-free equilibrium, (*S*^*\**^,*I*^*\**^) = (1, 0), and an endemic equilibrium 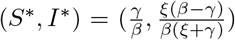.Since an epidemic requires that 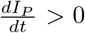 we substituted Equation 5 into this inequality and algebraically manipulated it until we obtained

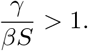

Assuming a completely susceptible population, or *S* = 1, we found the expression for the basic reproduction number for the system, 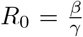.To assess the stability of this equilibrium, we formulated the Jacobian matrix

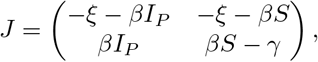

evaluated it at the disease-free equilibrium, and identified eigenvalues λ_1_ = *β*−*γ* and λ_2_ = −*ξ*. Because λ_2_ will always remain negative, values of λ_1_ determine stability. Notably, *β/γ >* 1 and *β* − *γ >* 0 are always satisfied simultaneously, meaning that the value of *R*_0_ determines equilibrium stability. These results are consistent with previous work [47, 48, 49] and provide a reference point for our more complex model that incorporates behavior, as well.

#### 3.1.2 Behavior model

To analytically characterize behavioral contagion dynamics in the absence of disease, we leverage model population characteristics 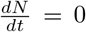 and *N* ≡ 1 to reduce the system outlined in Equations 6 and 7 to

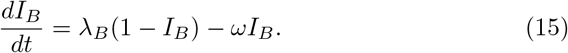

We then substituted in the previously mentioned expressions for λ_*B*_ and *M* to produce the expanded

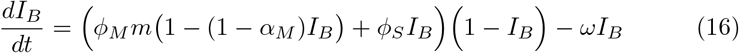

and identified two equilibrium solutions,

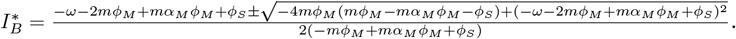

Though this equilibrium formula is not intuitive, it provides the initial impression that the baseline mosquito to human ratio *m* and other related parameters—namely, those weighting mosquito density _*M*_ and larval habitat reduction efficacy *α*_*M*_ —are influential to the behavior equilibrium. This makes sense given the dynamic feedback between the mosquito population and behavior, which are necessarily more difficult to disentangle than the one-way effect of social reinforcement. We also note that parameter values must satisfy the condition

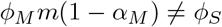

for these equilibria to exist. When this condition is violated, the right-hand side of Equation 16 no longer contains a quadratic dependence on *I*_*B*_, which changes the structure of the model to the point that these equilibria are no longer possible. Additionally, the parameter weighting the influence of mosquito density, ϕ _*M*_, must be positive for *I*_*B*_ to go from zero towards its non-zero equilibrium. When we examine Equation 16 with *ϕ*_*M*_ = 0, it simplifies to

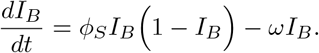

In a population without behavior already present (i.e., with *I*_*B*_ = 0), this reduced equation will always equal zero. We can interpret this to mean that in this model, mosquitoes must be a behavior stimulus for the behavior to arise in the first place.

We further explored model behavior and parameter relationships via linear stability analysis of Equation 16. Evaluating the first derivative of this equation at the positive equilibrium across a range of plausible parameters produces all negative values, indicating that these parameter combinations produce only stable solutions. The solutions associated with this analysis are shown in Figure 2, where we see that as the waning rate of the behavior decreases (i.e., the duration of the behavior increases), the equilibrium prevalence of behavior increases. In addition to the fact that this means that fewer individuals give up preventive behaviors in a given amount of time, more individuals in this category are gained as a result of a stronger influence of people taking up preventive behaviors due to social influence. We note that the sensitivity of 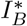 to *ϕ*_*S*_ becomes stronger in Figure 2 as *ω* decreases, which supports this interpretation.

**Figure 2:**
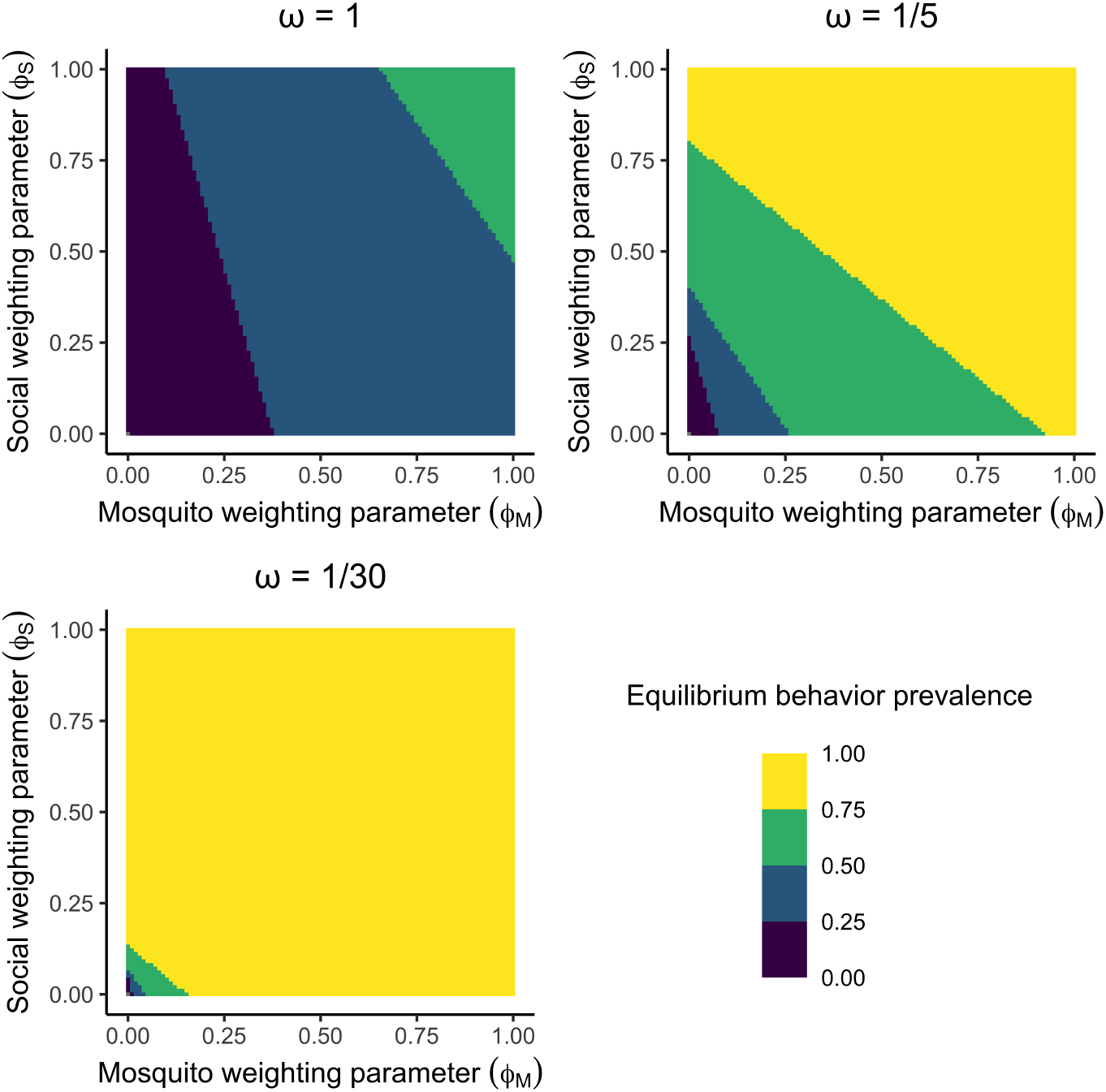
Prevalence of behavior under varying behavior waning rates *ω*. Fill color indicates the equilibrium prevalence of the behavior in a population. We assume the baseline value for mosquito control efficacy, *α*_*M*_ = 0.01, while varying weighting parameters for the mosquito population _*ϕM*_ and social pressure _*S*_.

### 3.2 Coupled contagion equilibria

We approached our analysis of the coupled contagion model numerically. Informed by our analysis of the single contagion model for behavior, we set all ϕ parameters to 0.01 to ensure that the magnitude of observed disease and behavior remained within reasonable bounds over the course of a simulation. Furthermore, setting the baseline mosquito-to-human ratio *m* to 1 ensured that the product *ϕ*_*M*_ *M* was equal to *ϕ*_*S*_ and *ϕ*_*D*_. These parameter choices established a baseline against which the effects of changes to parameters could be easily interpreted.

Simulating the model across a five-year period in the absence of preventive behaviors, we observed an equilibrium disease prevalence of 0.019, with a cumulative annual incidence of 1.0 for the final year of the simulation and an estimated *R*_0_ of 2.0 (Figure 3). These outcomes are plausible according to prior work on dengue epidemiology [36, 37] and were most sensitive to the tuned transmission parameters *b* and *c* (Table 1). When we introduced behavior into the model, we observed a reduction in equilibrium disease prevalence from 0.019 to 0.015, with a corresponding reduction in cumulative annual incidence from 1.0 to 0.8 and a reduction in *R*_0_ from 2.0 to 1.9, once equilibrium behavior prevalence was attained.

**Figure 3:**
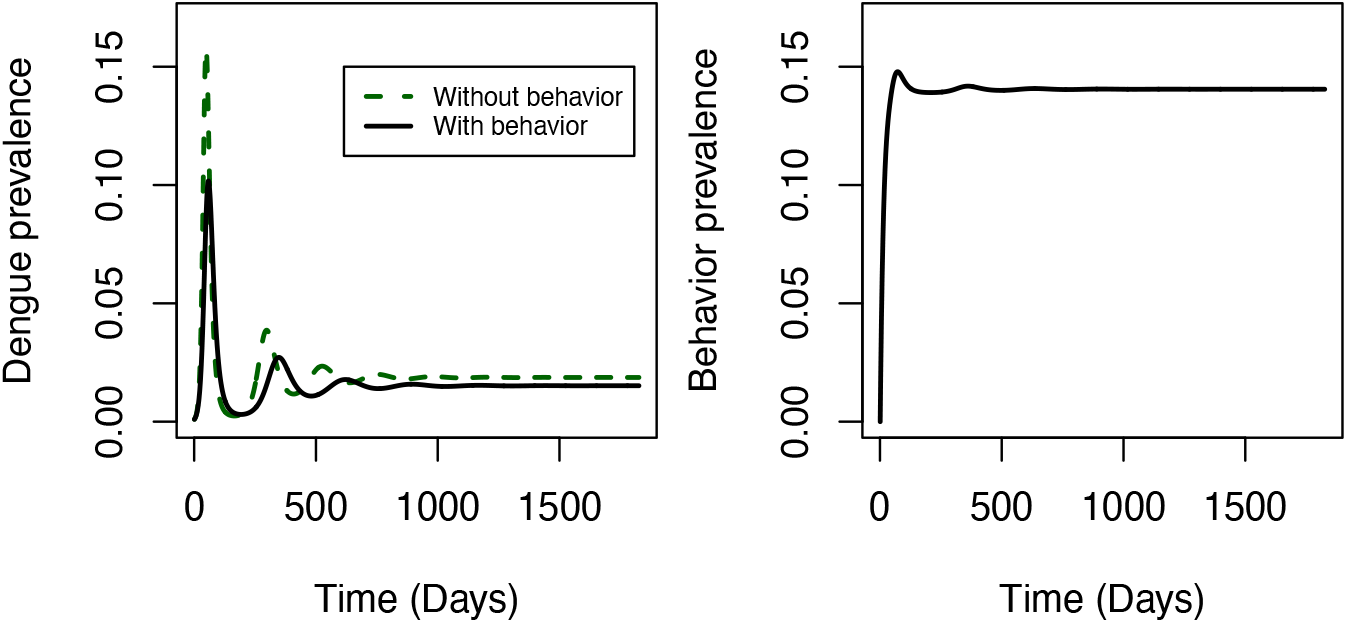
Disease and behavior dynamics over the course of a five year simulation. Line color and type indicate the presence or absence of the behavioral contagion.

To understand the relative influence of different behavioral parameters, we systematically removed each influence on behavior participation one by one under our baseline model parameterization (Figure 4). Removing the influence of disease (ϕ _*D*_ = 0) increased equilibrium disease prevalence only slightly, from 0.01517 to 0.01522. Removing the influence of social pressure (ϕ _*S*_ = 0) increased disease prevalence somewhat more, up to 0.160. Most significantly, removing the influence of mosquitoes (*ϕ*_*M*_ = 0) increased disease prevalence to 0.0186, which suggests that the influence of mosquitoes accounts for the majority of the reduction in disease prevalence when all influences on behavior are included. The extent of these reductions in disease correlate with the extent of change in the equilibrium behavior prevalence under each scenario about the parameters (Figure 4B). Ultimately, the relative importance of these influences on behavior owes to the frequency with which people susceptible to the behavior encounter them. The prevalence of disease is on the order of 1%, the prevalence of behavior is on the order of 10%, and the prevalence of people who are bitten by mosquitoes is 100%, consistent with the apparent strength of these influences in driving behavior participation.

**Figure 4:**
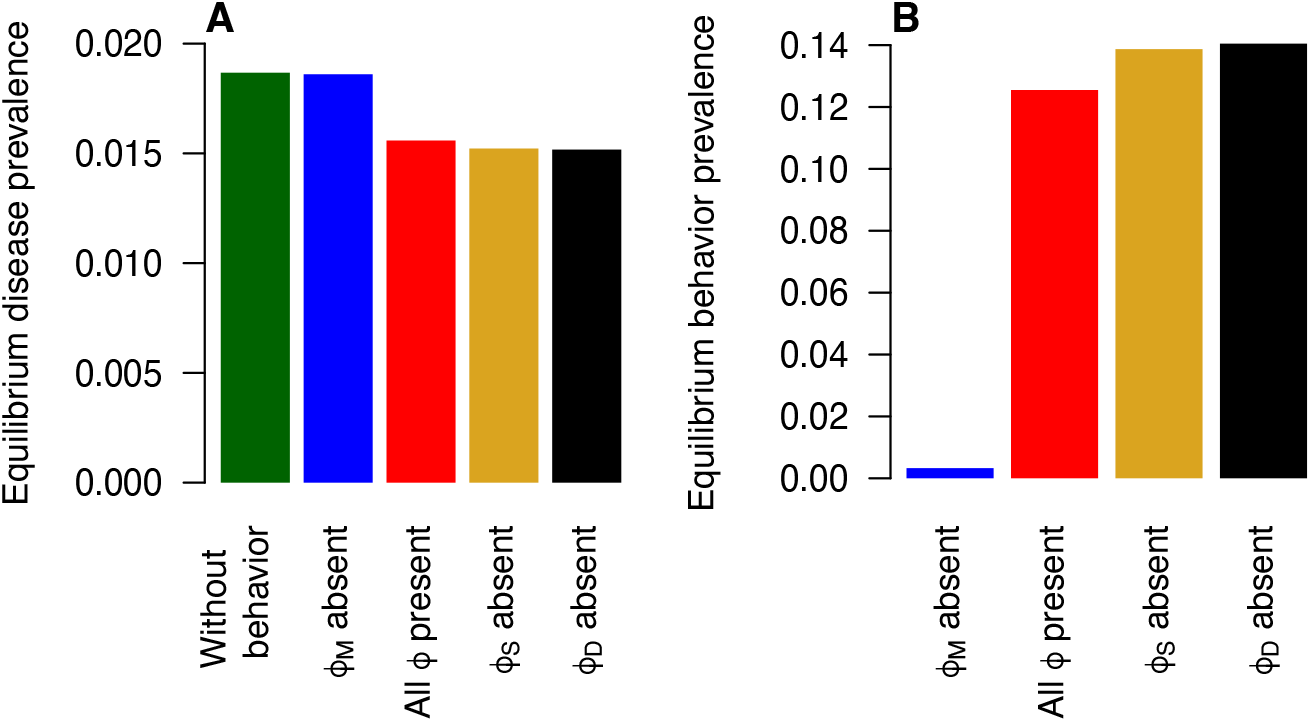
Equilibrium disease and behavior prevalence as influences on behavior participation are removed one by one. Fill color indicates the presence or absence of a given stimulus.

In general, we found that the effects of the ϕ parameters on disease were robust to values of other model parameters. Even so, the parameters governing the strength of the two forms of control played an important role in shaping these effects. First, changes in the relative risk of infection due to personal protection (*α*_*B*_) used by individuals in the behavior class had no effect on the prevalence of behavior but did affect disease prevalence (Figure 5). The curvilinear relationship between *α*_*B*_ and equilibrium disease prevalence reflects the quadratic effect of *α*_*B*_ on *R*_0_, given that it affects both mosquito bites required for a complete transmission cycle. Second, changes in the efficacy of community-level larval habitat reduction (1 *α*_*M*_) affected not only disease prevalence but also equilibrium behavior prevalence (Figure 6). This is a result of the fact that community-level control reduces mosquito density (*M*), which, in turn, reduces behavior prevalence. This reduction in behavior prevalence has the undesirable effect of reducing the use of personal protection. However, this undesirable effect is outweighed by the desirable effect of reducing mosquito density, resulting in a net reduction in disease prevalence.

**Figure 5:**
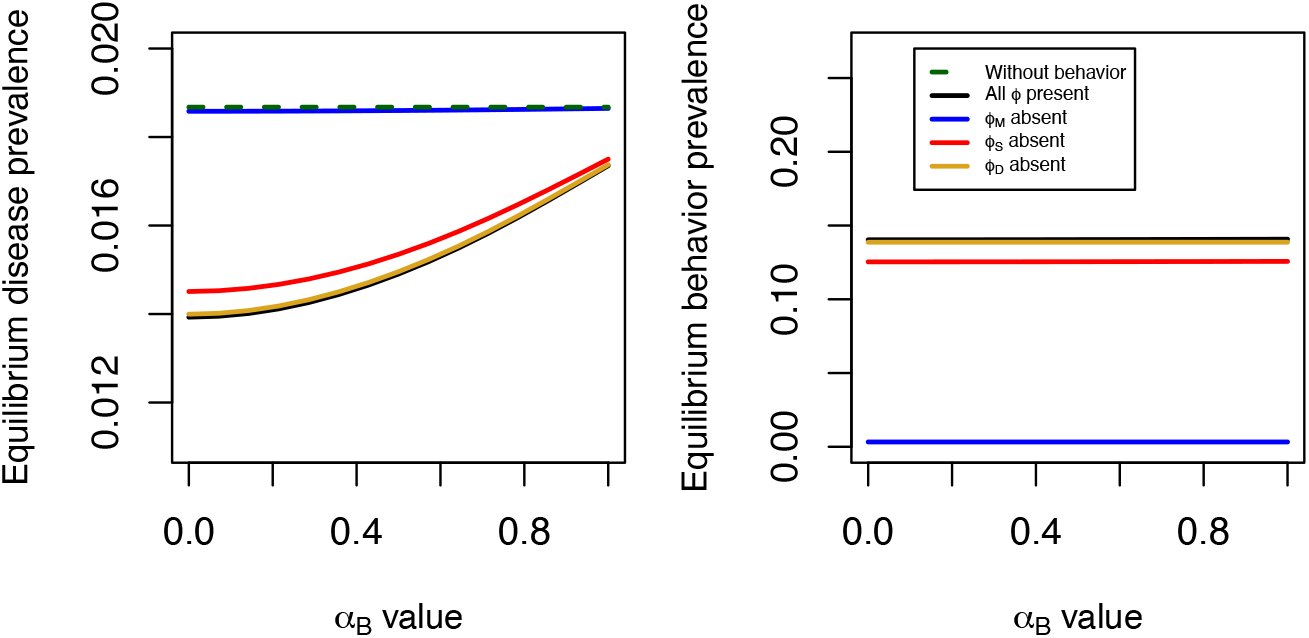
Equilibrium disease and behavior prevalence as relative risk associated with direct protection arising from preventive behavior *α*_*B*_ varies. Line colors indicate the behavior stimuli present or absent.

**Figure 6:**
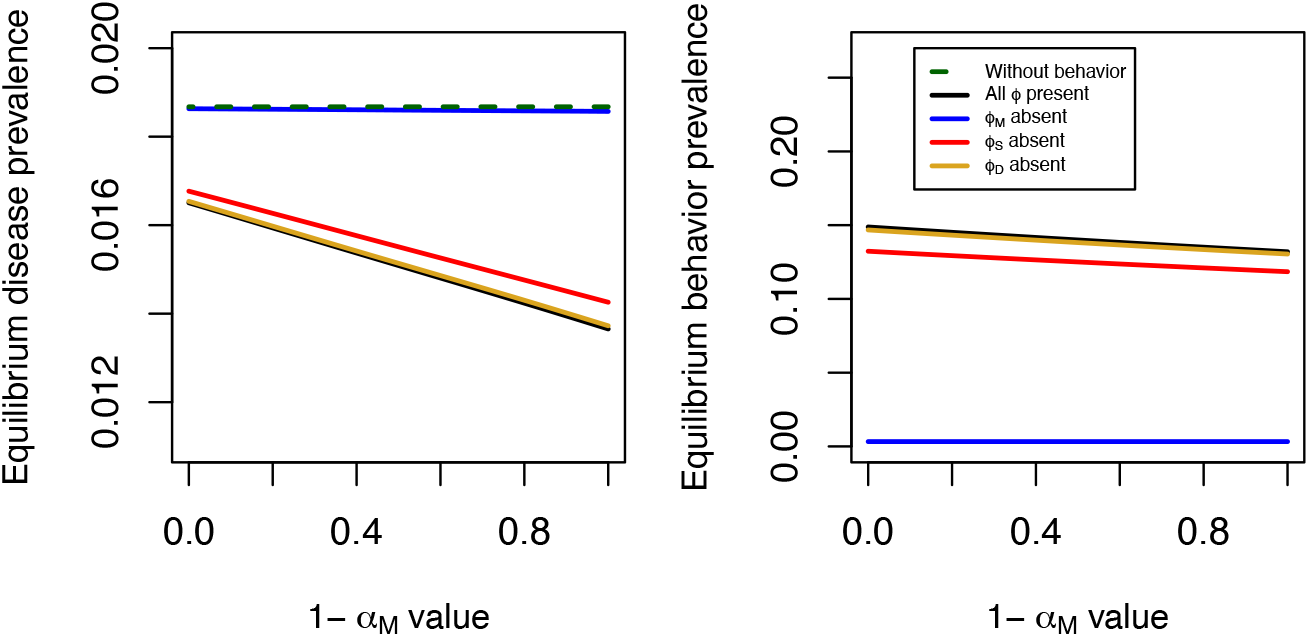
Equilibrium disease and behavior prevalence as the efficacy 1 *α*_*M*_ of community-level mosquito larval habitat reduction varies. Line colors indicate the behavior stimuli present or absent.

## 4 Discussion

In this study, we used a coupled contagion model of a mosquito-borne disease and mosquito preventive behaviors to gain insight into the primary drivers of feedbacks between disease and behavior for a mosquito-borne disease. We found that the coupling of disease and behavior contagions resulted in a lower equilibrium disease prevalence and a comparatively large equilibrium behavior prevalence. How long individuals maintain preventive behaviors was found to be a major driver of the equilibrium prevalence of behavior, which likewise affects the contribution of social pressure to behavior uptake. Interestingly, we found that the effects of contagion coupling on equilibrium prevalence of both contagions were most sensitive to the influence of mosquitoes in driving behavior uptake, followed by social pressure and, finally, disease itself. The relative importance of these influences reflects the frequency with which people encounter them. This finding highlights an important distinction between coupled contagion dynamics of mosquito-borne versus directly-transmitted disease systems, given that mosquitoes are not a relevant influence to the latter. Another feature unique to coupled contagion dynamics for mosquito-borne disease systems is that behaviors that reduce the mosquito population can also reduce behavior uptake due to reduced contact with (and annoyance by) mosquitoes. Counterintuitively, this can result in a diminishment of the benefits derived from reducing the mosquito population.

Beyond our study, epidemiological modeling studies that include preventive behaviors report measurable reductions in observed disease when such behaviors are introduced into the system [13, 50, 22, 21, 51]. This result is consistent with clinical trials and other field-based studies of community intervention performance [52, 7, 53]. Many of these modeling approaches include one or more parameters representing awareness of elements of the system (e.g., disease prevalence) as key drivers of participation in the behavior of interest. Increasing this awareness via a fixed parameter or via a weighting parameter for dynamic stimuli, as done here, is associated with more people engaging in behavior, regardless of specific behavioral mechanisms. At the same time, specific behavior- and disease-related outcomes are largely dependent on assumptions about stimulus strength and frequency. In contrast with directly-transmitted diseases, mosquito density adds complexity as both a stimulus and target of control, subject to both direct and indirect effects of preventive behaviors. While previous models of mosquito-borne disease and behavior do account for both community-level mosquito larval habitat reduction and individual-level personal protection, here we focus specifically on how they affect equilibrium prevalence of behavior and disease. In addition, we examined the effects of three distinct influence on the uptake of preventive behaviors. Our results reveal the consequences that even small changes to behavioral stimuli can have for equilibrium disease prevalence, highlighting the need for improved empirical understanding of these influences on the uptake of preventive behaviors.

In the absence of extensive empirical study, it can be difficult to explicitly quantify the attitudes and influences underpinning preventive behaviors. However, identifying the behavioral elements that drive effective mosquitoborne disease control — e.g., the success of interventions that incorporate community engagement and mobilization — could provide insight into a less-understood aspect of mosquito-borne disease control. Previous studies exploring community-based dengue interventions suggest that motivating factors are often time-varying [54], informed by socioeconomic and cultural expectations [55, 9], and shaped by inter- and intra-community dynamics [56, 57]. However, these factors are further complicated by the bidirectional feedback between disease and behavior, which makes it difficult to disentangle the two. There is a pressing need for more intentional field study development and data collection to provide insight into these unknowns [58, 12]. With such data, future refinement of the mechanisms driving the dynamics of preventive behaviors can be advanced [50].

In this work, we relied on simplifying assumptions to hone in on behavioral processes of interest and maintain analytical tractability in the single contagion transmission models. One such assumption was the use of a static mosquito-human ratio, rather than implementing one of the many alternative mosquito population model structures available [59, 60, 61]. There are also many other plausible approaches to modeling behavior that would address aspects of the behavior-disease relationship not featured in this study. First, our model assumes that individuals engaging in preventive behaviors stop doing so at a constant rate over time. Expanding our model to allow the three behavioral stimuli to influence not just the uptake of preventive behaviors, but also their continuation, would be one way to relax this assumption. Similarly, allowing for heterogeneity in behavior uptake and efficacy could address differences within and among groups noted in clinical trials of community-based mosquito interventions [62, 63]. Lastly, our identification of mosquito biting as the primary influence on behavior uptake could be sensitive to our model’s parameterization. Stimulus importance is context-dependent, though, and could be expected to vary under different conditions. For example, during an outbreak, people would likely find disease prevalence to be the most compelling influence towards control behaviors. Additionally, there could be a mismatch between the mosquitoes that influence preventive behaviors and those that pose a risk of infection [64].

Our work sought to characterize the interplay between disease and behavior in a general way for mosquito-borne diseases. We found that these diseases may be distinct from others given the wider range of stimuli that influence the uptake of preventive behaviors. We also found that there are distinct ways in which the use of personal protection and mosquito density reduction affect coupled contagion dynamics, with the latter showing an interesting feedback in which mosquito density reduction results in fewer mosquitoes, which in turn reduces the use of personal protection and contributes to more disease. In this situation, indirect protection and direct protection work against each other via changes in the prevalence of behavior, meaning that the relative efficacies of these interventions will dictate their net effect on disease in real-world systems. Likewise, empirical quantification of how the frequencies of encounters with the three behavioral stimuli we considered translate into preventive behavior uptake is an important priority for future work. Together, these findings highlight the importance of developing coupled contagion models of disease and behavior in disease systems with different transmission modes and other characteristics.

## Data Availability

Computer code is available from the authors upon request. Other than that, there are no data to make available.

## Acknowledgements

This material is based upon work supported by the National Science Foundation under Grant No. 2327814.

## References

[1] Sheng-Qun Deng et al. “A Review on Dengue Vaccine Development”. In: Vaccines 8.1 (Mar. 2020). Number: 1 Publisher: Multidisciplinary Digital Publishing Institute, p. 63. doi: 10.3390/vaccines8010063. url: https://www.mdpi.com/2076-393X/8/1/63 (visited on 09/21/2021).

[2] Senaka Rajapakse, Chaturaka Rodrigo, and Anoja Rajapakse. “Treatment of dengue fever”. In: Infection and Drug Resistance 5 (July 23, 2012), pp. 103–112. issn: 1178-6973. doi: 10.2147/IDR.S22613. url :https://www.ncbi.nlm.nih.gov/pmc/articles/PMC3411372/ (visited on 09/21/2021).

[3] Stephen J. Thomas, Daniel Strickman, and David W. Vaughn. “Dengue Epidemiology: Virus Epidemiology, Ecology, and Emergence”. In: Advances in Virus Research. Ed. by Thomas J. Chambers and Thomas P. Monath. Vol. 61. The Flaviviruses: Detection, Diagnosis, and Vaccine Development. Academic Press, Jan. 1, 2003, pp. 235–289. doi: 10.1016/S0065-3527(03)61006-7. url: https://www.sciencedirect.com/science/article/pii/S0065352703610067 (visited on 09/21/2021).

[4] Nicole L. Achee et al. “A Critical Assessment of Vector Control for Dengue Prevention”. In: PLOS Neglected Tropical Diseases 9.5 (May 7, 2015). Publisher: Public Library of Science, e0003655. issn: 1935-2735. doi: 10.1371/journal.pntd.0003655. url: https://journals.plos.org/plosntds/article?id=10.1371/journal.pntd.0003655 (visited on 09/22/2021).

[5] Daniel Pilger et al. “Dengue outbreak response: documented effective interventions and evidence gaps”. In: TropIKA.net 1.1 (Mar. 2010). Publisher: TDR/WHO. issn: 2078-8606. url: http://journal.tropika.net/scielo.php?script=sci_abstract&pid=S2078-86062010000100002&lng=en&nrm=iso&tlng=en (visited on 05/03/2023).

[6] Pablo A. Reyes-Castro et al. “Outdoor spatial spraying against dengue: A false sense of security among inhabitants of Hermosillo, Mexico”. In: PLOS Neglected Tropical Diseases 11.5 (May 17, 2017). Publisher: Public Library of Science, e0005611. issn: 1935-2735. doi: 10.1371/journal.pntd.0005611. url:https://journals.plos.org/plosntds/article?id=10.1371/journal.pntd.0005611 (visited on 05/03/2023).

[7] F. Espinoza-Gómez, C. Moises Hernández-Suárez, and R. Coll-Cárdenas. “Educational campaign versus malathion spraying for the control of Aedes aegypti in Colima, Mexico”. In: Journal of Epidemiology & Community Health 56.2 (Feb. 1, 2002). Publisher: BMJ Publishing Group Ltd Section: Public health policy and practice, pp. 148–152. issn: 0143-005X, 1470-2738. doi: 10.1136/jech.56.2.148. url:https://jech.bmj.com/content/56/2/148 (visited on 07/27/2021).

[8] Natarajan Arunachalam et al. “Community-based control of Aedes aegypti by adoption of eco-health methods in Chennai City, India”. In: Pathogens and Global Health 106.8 (Dec. 2012), pp. 488–496. issn: 2047-7732. doi: 10.1179/2047773212Y.0000000056.

[9] Céline Aerts et al. “Understanding the role of disease knowledge and risk perception in shaping preventive behavior for selected vector-borne diseases in Guyana”. In: PLoS Neglected Tropical Diseases 14.4 (Apr. 6, 2020), e0008149. issn: 1935-2727. doi: 10.1371/journal.pntd.0008149. url: https://www.ncbi.nlm.nih.gov/pmc/articles/PMC7170267/ (visited on 05/03/2023).

[10] Neil Andersson et al. “Evidence based community mobilization for dengue prevention in Nicaragua and Mexico (Camino Verde, the Green Way): cluster randomized controlled trial”. In: The BMJ 351 (July 8, 2015), h3267. issn: 0959-8138. doi: 10.1136/bmj.h3267. url: https://www.ncbi.nlm.nih.gov/pmc/articles/PMC4495677/ (visited on 09/21/2021).

[11] Jorge Arosteguí et al. “The Camino Verde intervention in Nicaragua, 2004–2012”. In: BMC Public Health 17 (Suppl 1 May 30, 2017), p. 406. issn: 1471-2458. doi: 10.1186/s12889-017-4299-3. url: https://www.ncbi.nlm.nih.gov/pmc/articles/PMC5506563/ (visited on 09/21/2021).

[12] Sebastian Funk, Marcel Salathé, and Vincent A. A. Jansen. “Modelling the influence of human behaviour on the spread of infectious diseases: a review”. In: Journal of The Royal Society Interface 7.50 (May 26, 2010). Publisher: Royal Society, pp. 1247–1256. doi:10.1098/rsif.2010.0142. url: https://royalsocietypublishing.org/doi/10.1098/rsif.2010.0142 (visited on 08/21/2024).

[13] Tyrus Berry et al. “Stabilizing the return to normal behavior in an epidemic”. In: medRxiv (Oct. 23, 2023), p. 2023.03.13.23287222. doi:10.1101/2023.03.13.23287222. url: https://www.ncbi.nlm.nih.gov/pmc/articles/PMC10055466/ (visited on 08/21/2024).

[14] S. Del Valle et al. “Effects of behavioral changes in a smallpox attack model”. In: Mathematical Biosciences 195.2 (June 1, 2005), pp. 228–251. issn: 0025-5564. doi:10.1016/j.mbs.2005.03.006. url: https://www.sciencedirect.com/science/article/pii/S0025556405000593 (visited on 08/21/2024).

[15] Matthew J Ferrari et al. “Network frailty and the geometry of herd immunity”. In: Proceedings of the Royal Society B: Biological Sciences 273.1602 (Nov. 7, 2006). Publisher: Royal Society, pp. 2743–2748. doi: 10.1098/rspb.2006.3636. url:https://royalsocietypublishing.org/doi/10.1098/rspb.2006.3636 (visited on 03/19/2021).

[16] Adaptive human behavior in epidemiological models. doi: 10.1073/pnas.1011250108. url: https://www.pnas.org/doi/10.1073/pnas.1011250108 (visited on 08/07/2024).

[17] Leah LeJeune et al. “Mathematical analysis of simple behavioral epidemic models”. In: Mathematical Biosciences 375 (Sept. 1, 2024), p. 109250. issn: 0025-5564. doi:10.1016/j.mbs.2024.109250. url: https://www.sciencedirect.com/science/article/pii/S002555642400110X (visited on 08/21/2024).

[18] Ceyhun Eksin, Keith Paarporn, and Joshua S. Weitz. “Systematic biases in disease forecasting – The role of behavior change”. In: Epidemics 27 (June 1, 2019), pp. 96–105. issn: 1755-4365. doi: 10.1016/j.epidem.2019.02.004. url:http://www.sciencedirect.com/science/article/pii/S1755436518301063 (visited on 10/22/2019).

[19] T. M. Q. R. Boccia et al. “Will people change their vector-control practices in the presence of an imperfect dengue vaccine?” In: Epidemiology & Infection 142.3 (Mar. 2014), pp. 625–633. issn: 0950-2688, 1469-4409. doi: 10.1017/S0950268813001350. url: https://www.cambridge.org/core/journals/epidemiology-and-infection/article/will-people-change-their-vectorcontrol-practices-in-the-presence-of-an-imperfect-dengue-vaccine/6A2ED05AD2D4615301F01F7D8E7ED148 (visited on 08/28/2024).

[20] Víctor Manuel Alvarado-Castro et al. “The influence of gender and temephos exposure on community participation in dengue prevention: a compartmental mathematical model”. In: BMC Infectious Diseases 24.1 (May 2, 2024), p. 463. issn: 1471-2334. doi: 10.1186/s12879-024-09341-w. url: https://doi.org/10.1186/s12879-024-09341-w (visited on 08/27/2024).

[21] Jing Jiao, Gonzalo P. Suarez, and Nina H. Fefferman. “How public reaction to disease information across scales and the impacts of vector control methods influence disease prevalence and control efficacy”. In: PLOS Computational Biology 17.6 (June 28, 2021). Publisher: Public Library of Science, e1008762. issn: 1553-7358. doi:10.1371/journal.pcbi.1008762. url:https://journals.plos.org/ploscompbiol/article?id=10.1371/journal.pcbi.1008762 (visited on 11/08/2021).

[22] Kimberlyn Roosa and Nina H. Fefferman. “A general modeling framework for exploring the impact of individual concern and personal protection on vector-borne disease dynamics”. In: Parasites & Vectors 15.1 (Oct. 8, 2022), p. 361. issn: 1756-3305. doi:10.1186/s13071-022-05481-7. url: https://doi.org/10.1186/s13071-022-05481-7 (visited on 05/03/2023).

[23] Joshua M. Epstein et al. “Coupled Contagion Dynamics of Fear and Disease: Mathematical and Computational Explorations”. In: PLOS ONE 3.12 (Dec. 16, 2008), e3955. issn: 1932-6203. doi:10.1371/journal.pone.0003955. url:https://journals.plos.org/plosone/article?id=10.1371/journal.pone.0003955 (visited on 11/29/2019).

[24] Kirti Jain et al. “Coupling Fear and Contagion for Modeling Epidemic Dynamics”. In: IEEE Transactions on Network Science and Engineering 10.1 (Jan. 2023). Conference Name: IEEE Transactions on Network Science and Engineering, pp. 20–34. issn: 2327-4697. doi:10.1109/TNSE.2022.3187775. url:https://ieeexplore.ieee.org/abstract/document/9813421?casa_token=WV3E_kJDYlgAAAAA:iU-4-W03U2o_9wIUFtBsJY6m06ovjabmhfkAkPespCJkxanXC3o4CzoCoMHConLQDHrKEMo (visited on 08/21/2024).

[25] Joshua M. Epstein, Erez Hatna, and Jennifer Crodelle. “Triple contagion: a two-fears epidemic model”. In: Journal of The Royal Society Interface 18.181 (Aug. 4, 2021). Publisher: Royal Society, p. 20210186. doi:10.1098/rsif.2021.0186. url:https://royalsocietypublishing.org/doi/full/10.1098/rsif.2021.0186 (visited on 08/21/2024).

[26] Nicola Perra et al. “Towards a Characterization of Behavior-Disease Models”. In: PLoS ONE 6.8 (Aug. 3, 2011). issn: 1932-6203. doi: 10.1371/journal.pone.0023084. url: https://www.ncbi.nlm.nih.gov/pmc/articles/PMC3149628/ (visited on 02/05/2020).

[27] Sansao A. Pedro et al. “Conditions for a Second Wave of COVID-19 Due to Interactions Between Disease Dynamics and Social Processes”. In: Frontiers in Physics 8 (2020). p. 428. issn: 2296-424X. doi: 10.3389/fphy.2020.574514. url: https://www.frontiersin.org/article/10.3389/fphy.2020.574514 (visited on 09/21/2021).

[28] Alejandro Bernardin, Alejandro J. Martínez, and Tomas Perez-Acle. “On the effectiveness of communication strategies as non-pharmaceutical interventions to tackle epidemics”. In: PLOS ONE 16.10 (Oct. 29, 2021). Publisher: Public Library of Science, e0257995. issn: 1932-6203. doi:10.1371/journal.pone.0257995. url: https://journals.plos.org/plosone/article?id=10.1371/journal.pone.0257995 (visited on 11/08/2021).

[29] Mathematica 14.0. Version 14.0.0.0. 2024. url:http://www.wolfram.com.

[30] Mikkel Meyer Andersen and Søren Højsgaard. caracas: Computer Algebra. Version 2.0.1. R package. 2023. url: https://github.com/rcas/caracas.

[31] Albert B. Sabin. “Research on Dengue during World War II”. In: The American Journal of Tropical Medicine and Hygiene 1.1 (Jan. 1, 1952). Publisher: The American Society of Tropical Medicine and Hygiene Section: The American Journal of Tropical Medicine and Hygiene, pp. 30–50. doi: 10.4269/ajtmh.1952.1.30. url: https://www.ajtmh.org/view/journals/tpmd/1/1/article-p30.xml (visited on 01/29/2025).

[32] Charlotte Probst et al. “The Normative Underpinnings of Population-Level Alcohol Use: An Individual-Level Simulation Model”. In: Health Education & Behavior 47.2 (Apr. 1, 2020). Publisher: SAGE Publications Inc, pp. 224–234. issn: 1090-1981. doi: 10.1177/1090198119880545. url:https://doi.org/10.1177/1090198119880545 (visited on 08/01/2022).

[33] Joshua M. Epstein. Agent Zero. Princeton University Press, Feb. 23, 2014. 272 p. isbn: 978-0-691-15888-4.

[34] Tuong Manh Vu et al. “Toward inverse generative social science using multi-objective genetic programming”. In: Proceedings of the Genetic and Evolutionary Computation Conference. GECCO ‘19: Genetic and Evolutionary Computation Conference. Prague Czech Republic: ACM, July 13, 2019, pp. 1356–1363. isbn: 978-1-4503-6111-8. doi: 10.1145/3321707.3321840. url:https://dl.acm.org/doi/10.1145/3321707.3321840 (visited on 08/11/2021).

[35] David L Smith et al. “Ross, Macdonald, and a Theory for the Dynamics and Control of Mosquito-Transmitted Pathogens”. In: PLoS Pathogens 8.4 (2012).

[36] Robert C. Reiner et al. “Time-varying, serotype-specific force of infection of dengue virus”. In: Proceedings of the National Academy of Sciences 111.26 (July 1, 2014). Publisher: National Academy of Sciences Section: PNAS Plus, E2694–E2702. issn: 0027-8424, 1091-6490. doi: 10.1073/pnas.1314933111. url: https://www.pnas.org/content/111/26/E2694 (visited on 09/24/2021).

[37] Lorenzo Cattarino et al. “Mapping global variation in dengue transmission intensity”. In: Science Translational Medicine 12.528 (Jan. 29, 2020). Publisher: American Association for the Advancement of Science, eaax4144. doi: 10.1126/scitranslmed.aax4144.url:https://www.science.org/doi/10.1126/scitranslmed.aax4144 (visited on 08/26/2024).

[38] Amy C. Morrison et al. “Efficacy of a spatial repellent for control of Aedes-borne virus transmission: A cluster-randomized trial in Iquitos, Peru”. In: Proceedings of the National Academy of Sciences 119.26 (June 28, 2022). Publisher: Proceedings of the National Academy of Sciences, e2118283119. doi: 10.1073/pnas.2118283119. url: https://www.pnas.org/doi/full/10.1073/pnas.2118283119 (visited on 08/21/2024).

[39] Helen J. Wearing and Pejman Rohani. “Ecological and immunological determinants of dengue epidemics”. In: Proceedings of the National Academy of Sciences 103.31 (Aug. 2006). Publisher: Proceedings of the National Academy of Sciences, pp. 11802–11807. doi: 10.1073/pnas.0602960103. url: https://www.pnas.org/doi/full/10.1073/pnas.0602960103 (visited on 08/26/2024).

[40] Nicholas G. Reich et al. “Interactions between serotypes of dengue highlight epidemiological impact of cross-immunity”. In: Journal of The Royal Society Interface 10.86 (Sept. 6, 2013), p. 20130414. issn: 1742-5689, 17425662. doi: 10.1098/rsif.2013.0414. url:https://royalsocietypublishing.org/doi/10.1098/rsif.2013.0414 (visited on 08/26/2024).

[41] Miranda Chan and Michael A. Johansson. “The Incubation Periods of Dengue Viruses”. In: PLoS ONE 7.11 (Nov. 30, 2012). issn: 1932-6203. doi:10.1371/journal.pone.0050972. url: https://www.ncbi.nlm.nih.gov/pmc/articles/PMC3511440/ (visited on 10/02/2020).

[42] Quirine A. Ten Bosch et al. “Community-level impacts of spatial repellents for control of diseases vectored by Aedes aegypti mosquitoes”. In: PLOS Computational Biology 16.9 (Sept. 25, 2020). Ed. by Jennifer A. Flegg, e1008190. issn: 1553-7358. doi: 10.1371/journal.pcbi.1008190. url:https://dx.plos.org/10.1371/journal.pcbi.1008190 (visited on 08/26/2024).

[43] Laura C. Harrington et al. “Analysis of Survival of Young and Old Aedes aegypti (Diptera: Culicidae) from Puerto Rico and Thailand”. In: Journal of Medical Entomology 38.4 (July 1, 2001), pp. 537–547. issn: 0022-2585, 1938-2928. doi:10.1603/0022-2585-38.4.537. url: https://academic.oup.com/jme/article-lookup/doi/10.1603/0022-2585-38.4.537 (visited on 08/26/2024).

[44] Thomas W. Scott et al. “Longitudinal Studies of Aedes aegypti (Diptera: Culicidae) in Thailand and Puerto Rico: Blood Feeding Frequency”. In: Journal of Medical Entomology 37.1 (Jan. 1, 2000), pp. 89–101. issn: 0022-2585. doi: 10.1603/0022-2585-37.1.89. url:https://doi.org/10.1603/0022-2585-37.1.89 (visited on 08/26/2024).

[45] R Core Team. R: A Language and Environment for Statistical Computing. R Foundation for Statistical Computing. Vienna, Austria, 2021. url:https://www.R-project.org/.

[46] Karline Soetaert, Thomas Petzoldt, and R. Woodrow Setzer. “Solving Differential Equations in R: Package deSolve”. In: Journal of Statistical Software 33.9 (2010). pp. 1–25. doi: 10.18637/jss.v033.i09.

[47] Roy M. Anderson and Robert M. May. Infectious Diseases of Humans: Dynamics and Control. Oxford, New York: Oxford University Press, Sept. 24, 1992. 766 pp. isbn: 978-0-19-854040-3.

[48] Maia Martcheva. “Introduction to Epidemic Modeling”. In: An Introduction to Mathematical Epidemiology. Ed. by Maia Martcheva. Boston, MA: Springer US, 2015, pp. 9–31. isbn: 978-1-4899-7612-3. doi:10.1007/978-1-4899-7612-3_2. url:https://doi.org/10.1007/978-1-4899-7612-3_2 (visited on 08/27/2024).

[49] Fred Brauer, Carlos Castillo-Chavez, and Zhilan Feng. “Endemic Disease Models”. In: Mathematical Models in Epidemiology 69 (June 25, 2019), pp. 63–116. doi:10.1007/978-1-4939-9828-9_3. url:https://www.ncbi.nlm.nih.gov/pmc/articles/PMC7316091/ (visited on 08/27/2024).

[50] MatthewRyanet al. “A behaviour and disease transmission model: incorporating the Health Belief Model for human behaviour into a simple transmission model”. In: ().

[51] Jaime Cascante-Vega et al. “How disease risk awareness modulates transmission: coupling infectious disease models with behavioural dynamics”. In: Royal Society Open Science 9.1 (Jan. 12, 2022). Publisher: Royal Society, p. 210803. doi: 10.1098/rsos.210803. url:https://royalsocietypublishing.org/doi/full/10.1098/rsos.210803 (visited on 05/03/2023).

[52] V Vanlerberghe et al. “Community involvement in dengue vector control: cluster randomised trial”. In: BMJ 338 (jun09 1 June 9, 2009), b1959–b1959. issn: 0959-8138, 1468-5833. doi: 10.1136/bmj.b1959. url: https://www.bmj.com/lookup/doi/10.1136/bmj.b1959 (visited on 07/26/2021).

[53] Juliana Quintero et al. “Effectiveness of an intervention for Aedes aegypti control scaled-up under an inter-sectoral approach in a Colombian city hyper-endemic for dengue virus”. In: PLOS ONE 15.4 (Apr. 1, 2020). Publisher: Public Library of Science, e0230486. issn: 1932-6203. doi: 10.1371/journal.pone.0230486.url: https://journals.plos.org/plosone/article?id=10.1371/journal.pone.0230486 (visited on 09/21/2021).

[54] Jocelyn Raude et al. “Understanding health behaviour changes in response to outbreaks: Findings from a longitudinal study of a large epidemic of mosquito-borne disease”. In: Social Science & Medicine 230 (June 1, 2019), pp. 184–193. issn: 0277-9536. doi: 10.1016/j.socscimed.2019.04.009. url: https://www.sciencedirect.com/science/article/pii/S0277953619302096 (visited on 10/12/2021).

[55] L. S. Lloyd et al. “The design of a community-based health education intervention for the control of Aedes aegypti”. In: The American Journal of Tropical Medicine and Hygiene 50.4 (Apr. 1994), pp. 401–411. issn: 0002-9637. doi: 10.4269/ajtmh.1994.50.401.

[56] Andrea Caprara et al. “Entomological impact and social participation in dengue control: a cluster randomized trial in Fortaleza, Brazil”. In: Transactions of the Royal Society of Tropical Medicine and Hygiene 109.2 (Feb. 2015), pp. 99–105. issn: 1878-3503. doi: 10.1093/trstmh/tru187.

[57] Alison M. Buttenheim et al. “Is participation contagious? Evidence from a household vector control campaign in urban Peru”. In: J Epidemiol Community Health 68.2 (Feb. 1, 2014). Publisher: BMJ Publishing Group Ltd Section: Research report, pp. 103–109. issn: 0143-005X, 1470-2738. doi:10.1136/jech-2013-202661.url: https://jech.bmj.com/content/68/2/103 (visited on 10/13/2021).

[58] Jamie Bedson et al. “A review and agenda for integrated disease models including social and behavioural factors”. In: Nature Human Behaviour 5.7 (Jul. 2021). Bandiera abtest: a Cg type: Nature Research Journals Number: 7 Primary atype: Reviews Publisher: Nature Publishing Group Subject term: Applied mathematics;Ecological epidemiology;Epidemiology;Health policy;Sociology Subject term id: applied-mathematics;ecological-epidemiology;epidemiology;healthpolicy;sociology, pp. 834–846. issn: 2397-3374. doi:10.1038/s41562-021-01136-2. url:http://www.nature.com/articles/s41562-021-01136-2 (visited on 10/12/2021).

[59] Krisztian Magori et al. “Skeeter Buster: A Stochastic, Spatially Explicit Modeling Tool for Studying Aedes aegypti Population Replacement and Population Suppression Strategies”. In: PLOS Neglected Tropical Diseases 3.9 (Sept. 1, 2009). Publisher: Public Library of Science, e508. issn: 1935-2735. doi: 10.1371/journal.pntd.0000508. url:https://journals.plos.org/plosntds/article?id=10.1371/journal.pntd.0000508 (visited on 08/21/2024).

[60] Emma L. Davis, T. Déirdre Hollingsworth, and Matt J. Keeling. “An analytically tractable, age-structured model of the impact of vector control on mosquito-transmitted infections”. In: PLOS Computational Biology 20.3 (Mar. 14, 2024). Publisher: Public Library of Science, e1011440. issn: 1553-7358. doi:10.1371/journal.pcbi.1011440. url: https://journals.plos.org/ploscompbiol/article?id=10.1371/journal.pcbi.1011440 (visited on 08/21/2024).

[61] M. Predescu et al. “On the dynamics of a deterministic and stochastic model for mosquito control”. In: Applied Mathematics Letters 20.8 (Aug. 1, 2007), pp. 919–925. issn: 0893-9659. doi: 10.1016/j.aml.2006.12.001. url: https://www.sciencedirect.com/science/article/pii/S0893965907000079 (visited on 08/27/2024).

[62] Jelte Elsinga et al. “Community participation in mosquito breeding site control: an interdisciplinary mixed methods study in Curaçao”. In: Parasites & Vectors 10.1 (Sept. 19, 2017), p. 434. issn: 1756-3305. doi:10.1186/s13071-017-2371-6. url:https://doi.org/10.1186/s13071-017-2371-6 (visited on 09/17/2021).

[63] Alidha Nur Rakhmani et al. “Factors associated with dengue prevention behaviour in Lowokwaru, Malang, Indonesia: a cross-sectional study”. In: BMC Public Health 18.1 (May 11, 2018), p. 619. issn: 1471-2458. doi:10.1186/s12889-018-5553-z. url: https://doi.org/10.1186/s12889-018-5553-z (visited on 08/27/2024).

[64] Andrew J. Mackay et al. “Dynamics of Aedes aegypti and Culex quinquefasciatus in Septic Tanks”. In: Journal of the American Mosquito Control Association 25.4 (Dec. 2009). Publisher: The American Mosquito Control Association, pp. 409–416. issn: 8756-971X, 1943-6270. doi:10.2987/09-5888.1. url:https://bioone.org/journals/journal-of-the-american-mosquito-control-association/volume-25/issue-4/09-5888.1/Dynamics-of-Aedes-aegypti-and-Culex-quinquefasciatus-in-Septic-Tanks/10.2987/09-5888.1.full (visited on 08/21/2024).

